# Integration of population-level data sources into an individual-level clinical prediction model for dengue virus test positivity

**DOI:** 10.1101/2023.08.08.23293840

**Authors:** RJ Williams, Ben J. Brintz, Gabriel Ribeiro Dos Santos, Angkana Huang, Darunee Buddhari, Surachai Kaewhiran, Sopon Iamsirithaworn, Alan L. Rothman, Stephen Thomas, Aaron Farmer, Stefan Fernandez, Derek A T Cummings, Kathryn B Anderson, Henrik Salje, Daniel T. Leung

## Abstract

The differentiation of dengue virus (DENV) infection, a major cause of acute febrile illness in tropical regions, from other etiologies, may help prioritize laboratory testing and limit the inappropriate use of antibiotics. While traditional clinical prediction models focus on individual patient-level parameters, we hypothesize that for infectious diseases, population-level data sources may improve predictive ability. To create a clinical prediction model that integrates patient-extrinsic data for identifying DENV among febrile patients presenting to a hospital in Thailand, we fit random forest classifiers combining clinical data with climate and population-level epidemiologic data. In cross validation, compared to a parsimonious model with the top clinical predictors, a model with the addition of climate data, reconstructed susceptibility estimates, force of infection estimates, and a recent case clustering metric, significantly improved model performance.

## Introduction

Acute febrile illness (AFI) is a common reason for seeking healthcare in low- and middle-income countries (LMICs) (*1*). Determination of AFI etiology is often limited by diagnostic testing capacity, given the wide spectrum of potential infectious agents. Inappropriate use of testing and treatment resources may result in poor outcomes, such as the high case fatality rates seen in admitted AFI patients (5-20%) (*2-7*). Dengue virus (DENV) is a major cause of AFI in LMICs, accounting for an estimated 390 million infections, 96 million illnesses, 2 million severe cases, and 21,000 deaths per year (*8*). The differentiation between dengue and other common causes of febrile illness is important to avoid misdiagnosis, which can lead to delays in initiation of effective treatment, and inappropriate use of antibiotics (*9*). Due to the lack of pathognomonic clinical features that reliably distinguish dengue from other febrile illnesses, virological or serological laboratory confirmation is required for definitive diagnosis. While multiplexed tests that can quickly identify the causative pathogen are ideal, they are often unavailable in LMICs due to cost and insufficient laboratory infrastructure. Even rapid, point-of-care tests may be cost-prohibitive in LMICs (*10*). Accurate and cost-effective tools to better determine etiology of fever at the point-of-care are greatly needed to guide the use of diagnostics and therapeutics, conserving scarce healthcare resources.

Clinical Decision-Support Systems (CDSS) incorporating prediction models may offer a solution to better management of infectious diseases in low resource settings. CDSSs, such as applications on smartphone devices, can gather data from a range of online sources and implement sophisticated clinical prediction models that would be impractical for clinicians to calculate manually. CDSS have proven effective at improving therapeutic management and reducing unnecessary diagnostic tests in both high-income countries (HICs) (*11*) and LMICs (*12-14*). In Bangladesh, an electronic CDSS was shown to improve clinical dehydration assessment and WHO diarrhea guideline adherence, as well as reduce non-indicated antibiotic use in children under five by 29% (*12*). Traditional predictive models generally incorporate clinical information that is obtained solely from the presenting patient. Predictive models that incorporate additional information – such as seasonal or climate predictors, location-specific historical prevalence, characteristics of prior patients – have been shown to increase diagnostic accuracy and limit inappropriate antibiotic use (*14-16*).

The underlying probability of being infected by DENV varies by both space and time. The risk of DENV transmission depends on conditions that promote mosquito breeding, including when temperatures are warmer (*17-19*), and the risk of infection is influenced by local population immunity, as large outbreak years are typically followed by periods of low transmission (*20-22*). As most DENV transmission is highly focal, it means that population susceptibility profiles can be spatially heterogeneous at any time (*21, 23-25*). Thus, our objective is to develop an improved clinical prediction model for dengue by integrating temporal and spatial (location-specific) parameters including climate data, clustering of recent cases, and population susceptibility estimates derived from seroprevalence or hospital data in the surrounding community. We demonstrate the potential for integrating location- and population-specific data sources into clinical prediction models, with the potential to inform the development of improved tools to aid clinicians in diagnostic and therapeutic decision making for patients presenting with suspected dengue.

## Methods

### Location

Kamphaeng Phet is a province in north-central Thailand that is located 350 km north of Bangkok and has a population of 725,000 people in a mostly rural and semirural setting (*26, 27*). We used data collected from patients presenting to Kamphaeng Phet Provincial Hospital (KPPH), a large, tertiary care hospital in the province to identify clinical predictors that could discriminate between DENV-infected and uninfected patients (*26, 27*).

### Hospital-based suspected dengue patient data

We used data on over 12,000 patients presenting to KPPH with suspected dengue between August 2007-December 2021. The data was collected by the United States Army Medical Directorate-Armed Forces Research Institute of Medical Sciences (USAMD-AFRIMS). As DENV testing in this hospital is provided free of charge and this is a highly DENV-endemic region, individuals will be tested for DENV infection if there is any suspicion of dengue, however minor. This provides an excellent test case to understand whether individual or location-specific risk factors are associated with testing positive for DENV.

For all suspected dengue cases, we used demographic and clinical information including patient age, sex, home village, admission diagnosis, date of admission, presenting symptoms, and DENV PCR status. The following signs and symptom were recorded as binary variables: fever, chills, malaise, rhinitis, rash, sore throat, seizure, cough, nuchal rigidity, eye pain, nausea, headaches, vomiting, joint pain, abnormal movements, anorexia, myalgias, diarrhea, dark urine, abdominal pain, and bleeding. DENV infection was evaluated using RT-PCR. We recorded the residence of each patient to the district (Amphoe) level using detailed base maps of the region.

### Climate variables using National Oceanic and Atmospheric Administration (NOAA) data

Climate and seasonal factors such as temperature, precipitation, and humidity influence vector populations and DENV transmission (*17-19, 28*). We employed the R package GSODR to gather climate data from the central most NOAA weather station in the province of Kamphaeng Phet, Thailand, which included mean daily temperature, precipitation, dewpoint, relative humidity, sea level pressure, visibility, and windspeed. To better reflect seasonal trends, we aggregated data in 14-day increments prior to the day of the DENV infection prediction. As climate can alter vector feeding behavior (*19, 29*), we used aggregated climate predictors in the two weeks prior to case presentation. Additionally, climate in the months prior to outbreaks can influence both vector population dynamics as well as viral replication (*19, 28*). To determine the appropriate lag time for each climate variable, we constructed a random forest classifier with climate variables lagged at one, two, and three months. Using the R package, “vip”, we calculated each Variable of Importance by AUC and used the best performing lag time for each climate variable.

### Estimates of temporal changes in population susceptibility using national surveillance system data

We estimate population susceptibility data using age-specific case data from the national surveillance system using data from Kamphaeng Phet province only. We note that most of the cases in this dataset are suspected DENV cases (i.e., without confirmatory testing). We have previously developed models to explicitly link underlying infection risks to the observed age distribution of cases by age and year to estimate annual age-specific force of infection in provinces of Thailand up until 2017 (*30*). The estimates can be used to reconstruct the buildup of immunity in populations by age. Here, we reconstruct population susceptibilities in Kamphaeng Phet going into each year, using only data prior to the year, to mimic the real-world use, where only prior years’ data is available. As dengue disease severity is greatest for secondary infections, we consider two alternative formulations to define susceptibility to disease. Firstly, we consider complete susceptibility, where we use the estimates of the proportion of individuals of an age group and year that are completely seronaive. Second, we consider the proportion of individuals of an age group and year that have experience one prior infection, and are therefore at risk of increased risk of severe disease.

### Estimates of spatial differences in the underlying force of infection using seroprevalence data from a cohort study

To estimate underlying spatial differences in the force of infection in the province, we make use of a DENV cohort study in the region, where healthy individuals of all ages from throughout Kamphaeng Phet province have provided blood (*31*). The cohort is ongoing. We use data from samples collected during baseline blood draws, that occurred between 2015 and 2021. Hemagglutination inhibition assays were used to characterize immunity to the four DENV serotypes; individuals were considered seropositive if they had a titer of 10 or greater to any serotype. We have previously used this seroprevalence data to estimate the underlying mean force of infection, and the proportion of the population that are susceptible to DENV infection in different subdistricts in the province (*32*). Here, we use this subdistrict specific estimates to characterize underlying heterogeneity in the force of infection in the province. As the cohort data comes from 2015-2021, however, much of the hospital case data we are working with comes from prior to the cohort, we are assuming that the force of infection is stable in time within any location.

### Spatial clustering of positive cases based on prior patients presenting to the hospital

The local clustering of positive cases from a single area, may signal local ongoing transmission. To assess for a temporal and spatial relationship between cases, we stratified cases that presented to KPP hospital by both district and province and then summed the number of positive cases in the 30 days prior to presentation divided by the total cases over the study period from that area.

### Statistical Analysis and Modeling

We fit random forest classifiers to predict DENV infection. Random forests are a machine learning method which constructs a multitude of decision trees and averages over them to obtain a prediction robust to nonlinearities and interactions between covariates, and has been widely applied to biomedical sciences for both classification and regression (*33, 34*).

We initially identified the subset of clinical symptoms that were most informative of true infection status. To do this we fit random forest models using only clinical predictors and then used the R package “vip” to calculate the Variable of Importance by AUC for each clinical variable. We determined a variable’s importance by calculating the change in AUC after permuting, or randomly shuffling each predictor. To attempt to achieve the most parsimonious prediction rule (i.e., the best predictive model requiring the fewest variables to be input by clinicians), we fit random forest and logistic regression models using training data with consecutively increasing clinical predictor set sizes based on the order of importance and applied this to the test set to determine the smallest model with the best performance. Next, we incorporated the patient extrinsic factors. We fit each random forest classifier using 1000 decision trees and used the default number of variables to be randomly considered at each node split (*mtry* = square root of number of candidate variables). In the construction of our predictive models, we input climate predictors, age, susceptibility estimates, and the case clustering metric as continuous variables and we input the optimized clinical predictors as binary presence or absence categorical variables. Missing predictor data was imputed using the R package ‘RandomForest’.

We used logistic regression for each predictor to create a univariate comparison between DENV-positive and DENV-negative cases. We fit multiple logistic regression models to compare the performance of parsimonious models with a random forest classifier using the same number of predictors.

To assess predictive performance for both random forest and logistic regression models, we used repeated cross-validation using 80% training/20% testing splits with 100 iterations. No testing data was used when training the model. In each iteration, predictions on the test set were produced and corresponding measures of performance obtained. To determine overall model performance, we averaged the area under the receiver operator characteristic curve (AUC) and confidence intervals for the 100 iterations. To determine statistical significance between models we used a bootstrap method over 100 iterations, which involves resampling the data with replacement multiple times, creating bootstrap samples. For each bootstrap sample, receiver operating characteristic (ROC) curves were generated and the differences between the curves were computed. All analyses were completed using R version 4.2.0, and model development/validation was completed in accordance with the TRIPOD checklist (Supplement Table S1).

### Ethical considerations

This study was approved by the institutional review boards of the Thai Ministry of Public Health and Walter Reed Army Institute of Research (WRAIR #2119), and the University of Utah (IRB_00150106)

## Results

Of the 12,833 participants in the clinical data set, 5731 (45%) were confirmed to have DENV infection by PCR. DENV-positive patients were significantly younger (18 vs 22 years, p<0.001, Table 1). Nearly all cases (97.8%) came from the 11 districts within Kamphaeng Phet province (Table 1). There was no significant difference between the probability of testing positive for males and females (p=0.07); no other genders were reported. The probability of testing positive differed substantially by age, ranging from 26% for those < 4 years to 58% for those 15-19 years of age (Table 2). Patients between the ages of 10-14 years, 15-19 years, and 5-9 years comprised the largest proportion of cases (23%, 18%, 16% respectively) while older patients comprised a much smaller proportion of cases (30-34 years 5%, 35-39 years 4%).

**Table 1:**
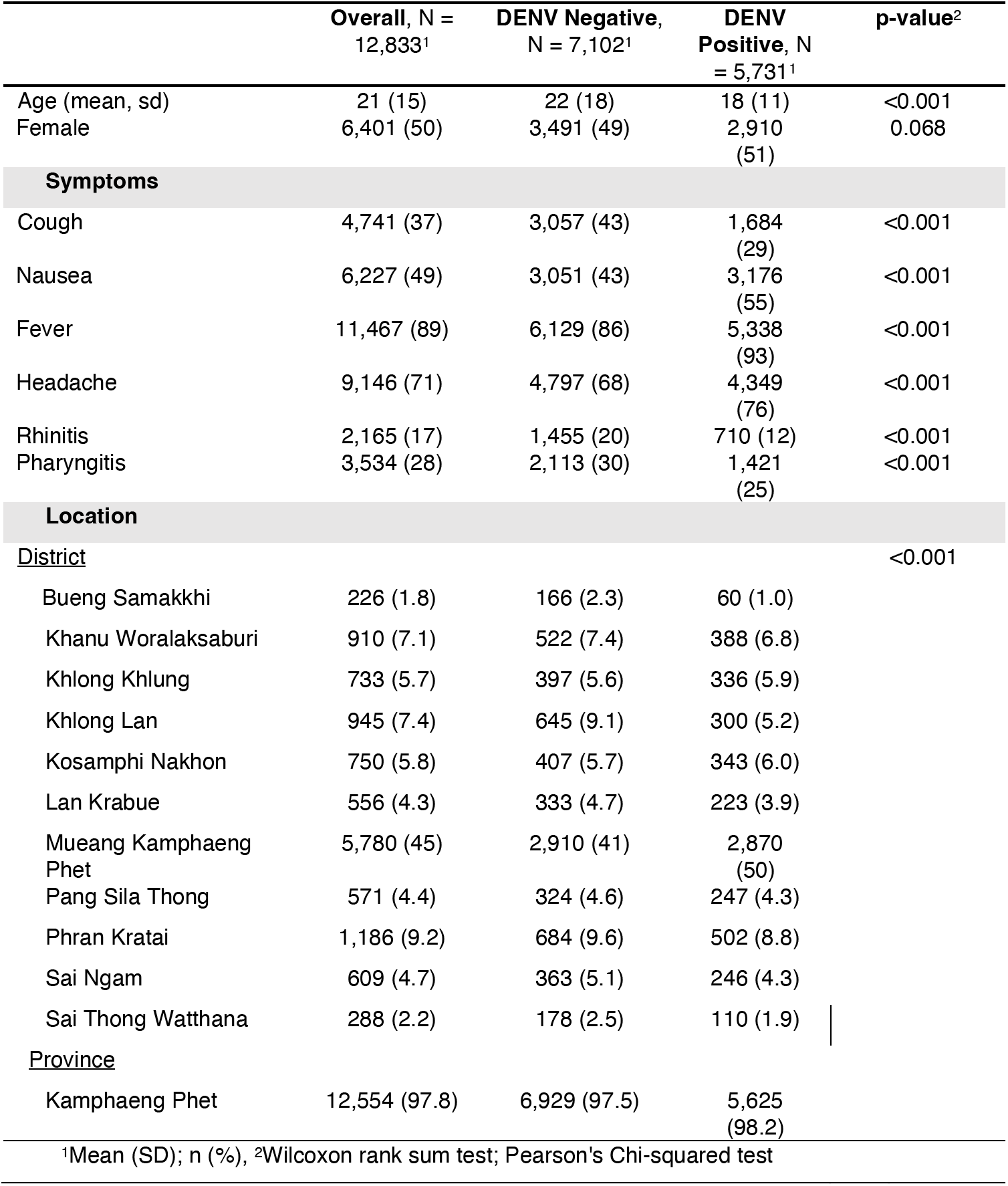
Age, gender, and top discriminative symptoms by DENV positivity. Locations listed are the eleven provinces in Kamphaeng Phet.

**Table 2.**
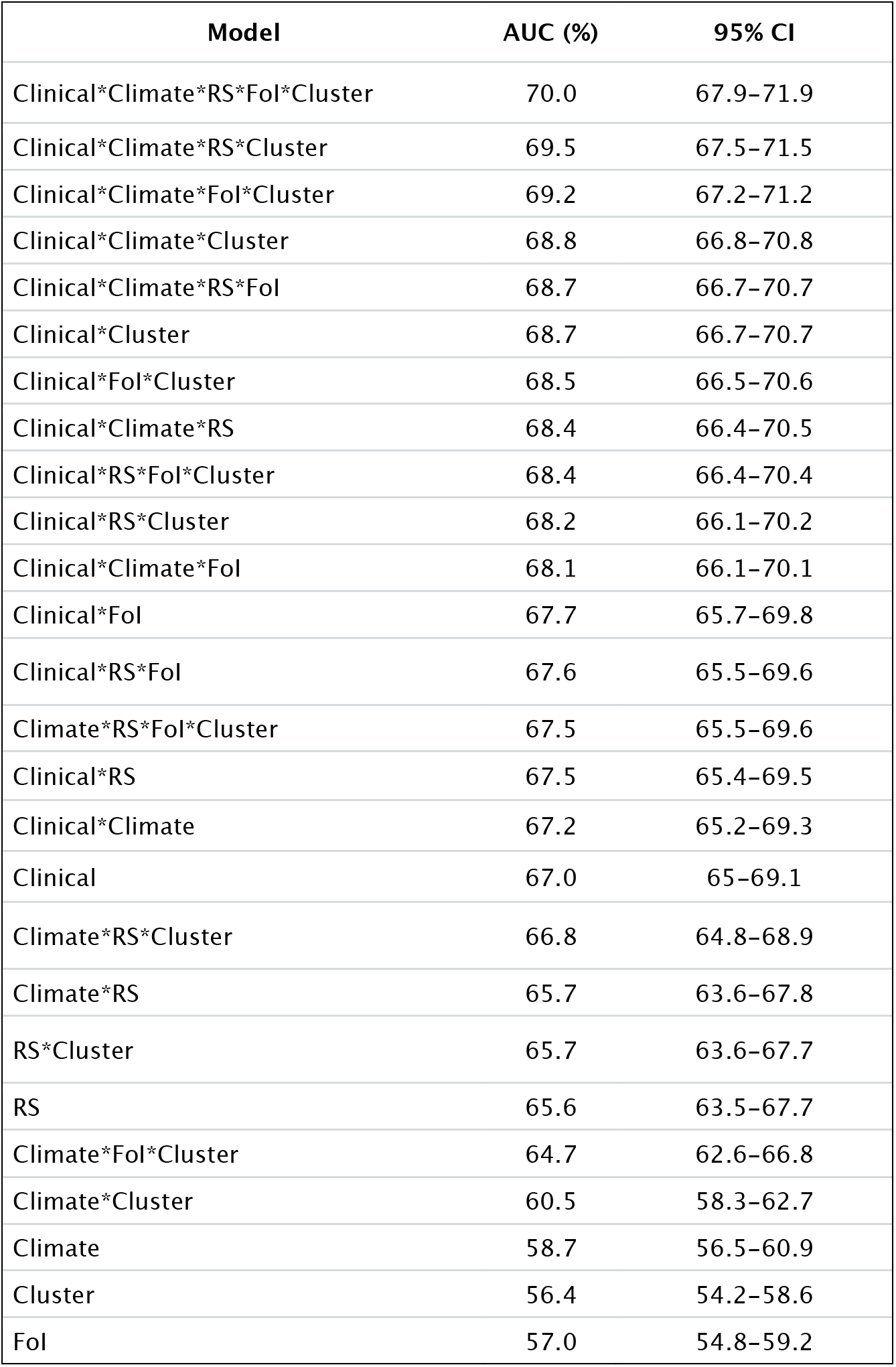
The AUCs and confidence intervals by base model, compared to base model plus inclusion of additional data sources. ‘Clinical’ indicates the inclusion of the top three clinical predictors, ‘Climate’ indicates the inclusion of climate predictors, ‘RS’ indicates the inclusion of reconstructed susceptibility estimates derived using national surveillance data, ‘FOI’ indicates the inclusion of force of infection estimates derived using cohort data, ‘Cluster’ indicates the recent case cluster metric.

We found that there were significant differences in the clinical symptoms between DENV positive and negative patients. Table 1 lists the top discriminative symptoms between the groups based on random forest and logistic regression. The most common symptom reported was fever, followed by headache. In univariate analysis, we found that individuals with fever, chills, malaise, retro-orbital pain, nausea, headache, and vomiting were significantly more likely to test positive for DENV, and individuals with cough, rhinitis, pharyngitis were significantly less likely to test positive for DENV (Supplementary Table S2).

When we examined the proportion of positive cases to total cases by year and month, we found that both total and positive cases significantly increased in the months between June and September (p<0.001). The proportion of positive cases differed substantially by year (p< 0.001), ranging from 19% in 2016 to 90% in 2017. The period of lowest test-positivity in 2016 and 2017, coincided with the Zika virus epidemic in the country (Figure 1).

**Figure 1.**
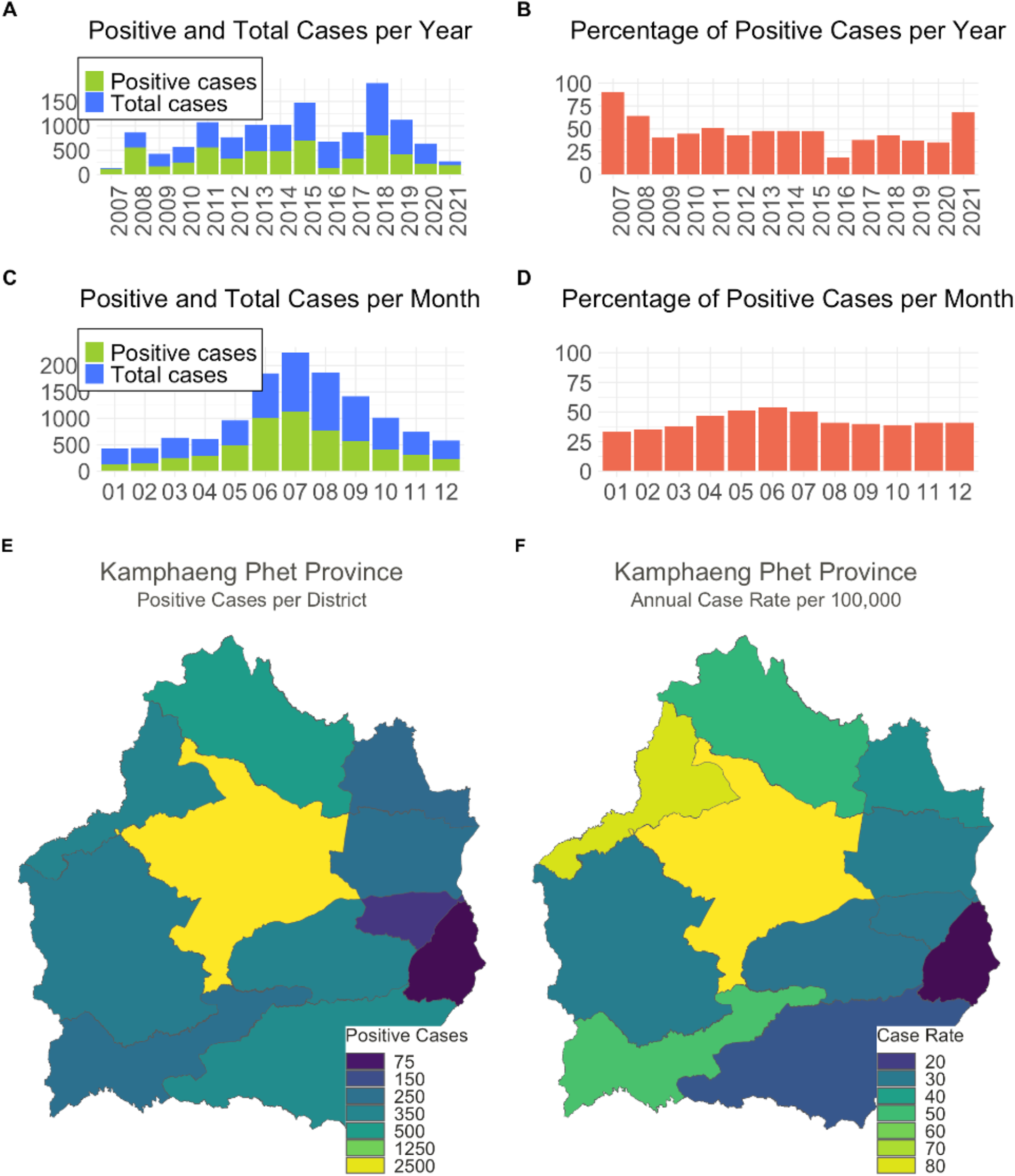
Dengue virus (DENV) cases at Kamphaeng Phet Provincial Hospital, Thailand, 2007-2021. The number of DENV cases (green) over total cases (blue) as proportion of AFI cases by year (A) and month (C) and the percentage of positive cases by year (B) and month (D) over the study period. A map of Kamphaeng Phet Province and its 11 districts. Colors indicate the number of positive cases (E) and the annual case rate per 100,000 persons (F) within each district between 2007-2021.

### Derivation of a model using clinical parameters alone resulted in a parsimonious model that achieved moderate predictive performance

We first assessed the performance of the model using a traditional clinical prediction model which only includes the presenting patient’s information. A random forest classifier using all 23 clinical features resulted in an average AUC of 69.5% (95%CI: 67.5-71.5) from repeated cross-validation. To determine the optimal number of variables for a parsimonious prediction model, we used a random forest classifier to analyze the improvement in model performance with each additional clinical variable included. Figure 2 shows the improvement in AUC with each additional variable using two random forest classifiers – one with all other predictors and the other using only clinical data – as well as a logistic regression model using only clinical variables. Performance levelled off with three clinical variables: age, cough, and nausea. Using a model with only these three predictors, we achieve an average AUC of 67.0% (95%CI: 65.0-69.1). Supplementary Table S3 shows the relative frequency of these variables by age group. We demonstrate the direction and magnitude of the effect of the top predictors by generating partial dependence plots from random forest and logistic regression classifiers (Supplementary Figure S1).

**Figure 2.**
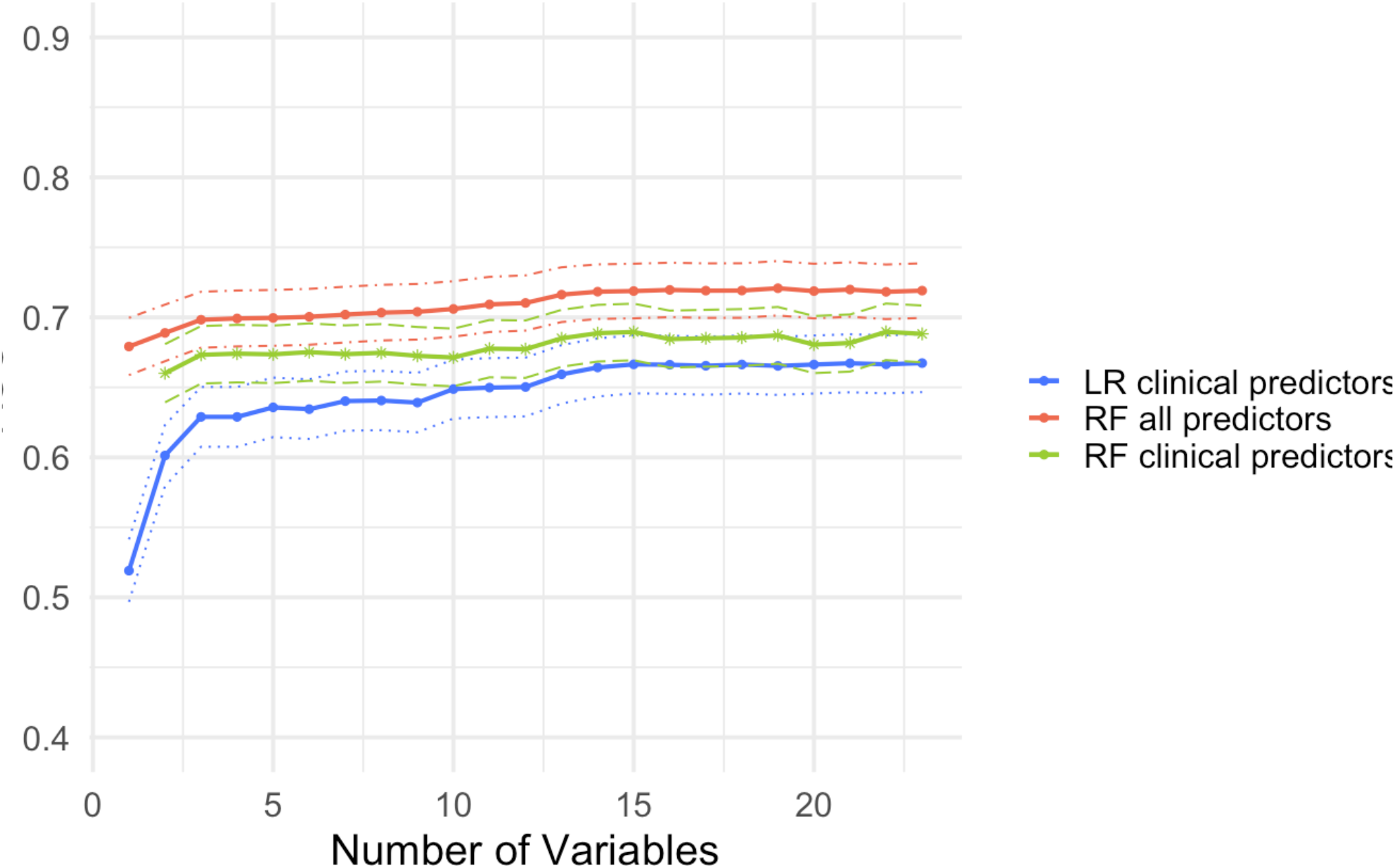
Average AUC and 95% CIs from cross-validation (100 iterations) for Random Forest (RF) and Logistic Regression (LR) models. The red line indicates an RF model with all other predictors (climate, reconstructed susceptibilities estimates, force of infection estimates, prior patients) included. The green line indicates an RF model which includes only clinical predictors. The blue line indicates an LR model with only clinical predictors included. The dotted lines indicate CIs.

### Addition of climate data to the clinical parameters model resulted in an improved area under the curve

Next, we fit models using climate data. To appropriately adjust lag time for each climate variable, we fit a random forest classifier using only climate variables and assessed the Variables of Importance by AUC. A random forest model with recent and lagged aggregated climate data without clinical predictors resulted in an AUC of 58.7% (95% CI: 56.5-60.9). We found the best performing climate variables were visibility, relative humidity, wind speed, and precipitation, all lagged by 3 months. For each climate predictor, Supplementary Table S4 lists the odds ratio and compares the mean of each predictor by DENV-positive or negative groups. Figure 3 shows the relationship between visibility, relative humidity, and the proportion of positive cases each month. When combined with the top three clinical variables, climate data performed similarly (median p = 0.60, 2% p-values <0.05) as clinical data alone. Table 2 shows the AUCs for the clinical base model, compared to the base model plus the inclusion of additional data sources.

**Figure 3.**
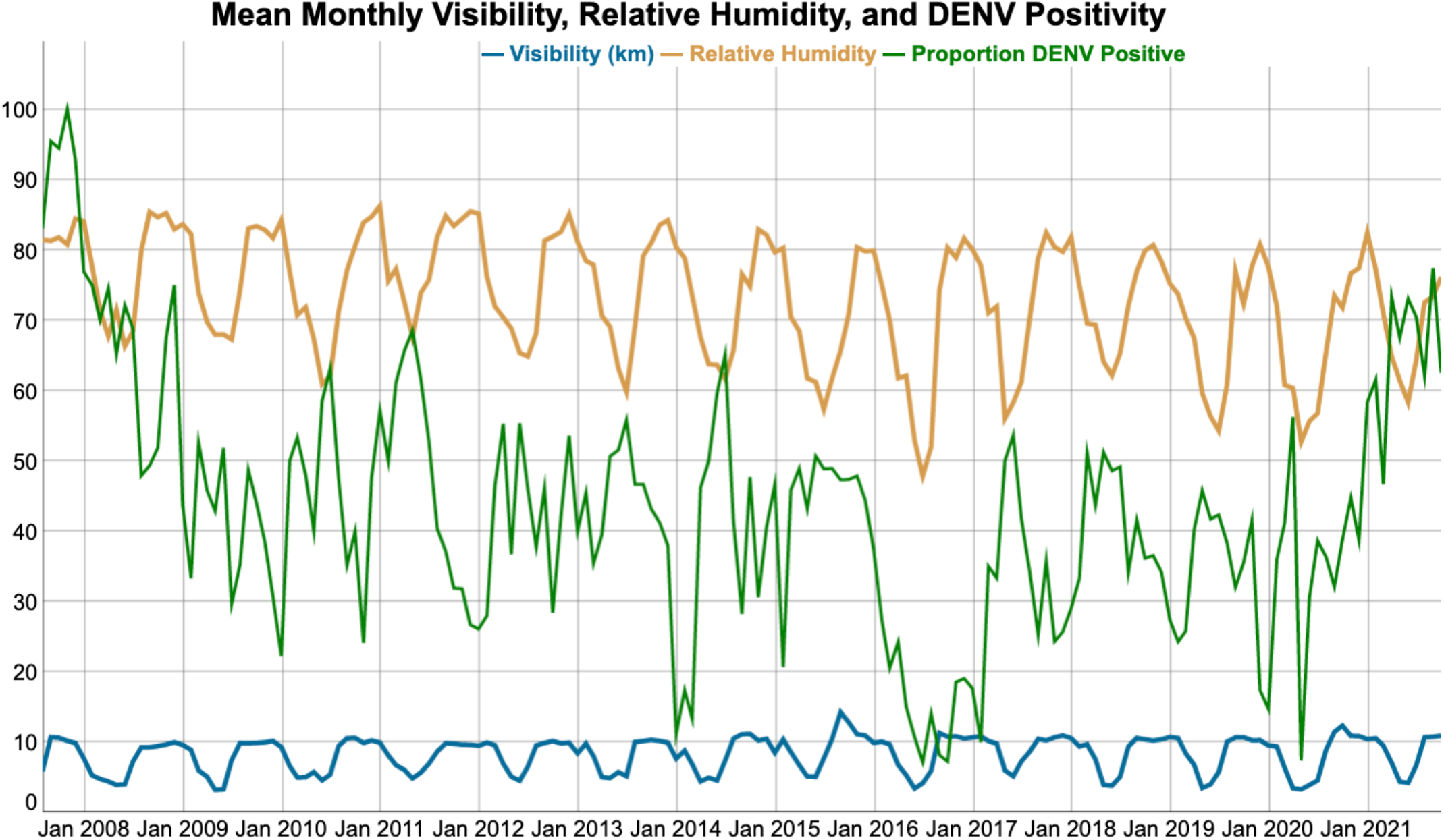
The monthly relative humidity (orange) and visibility (blue) in Thailand over the study period, compared with rates of DENV (green). For each case, we gathered the nearest NOAA weather station’s climate data, lagged by three months, and averaged that data for each month.

**Figure 4.**
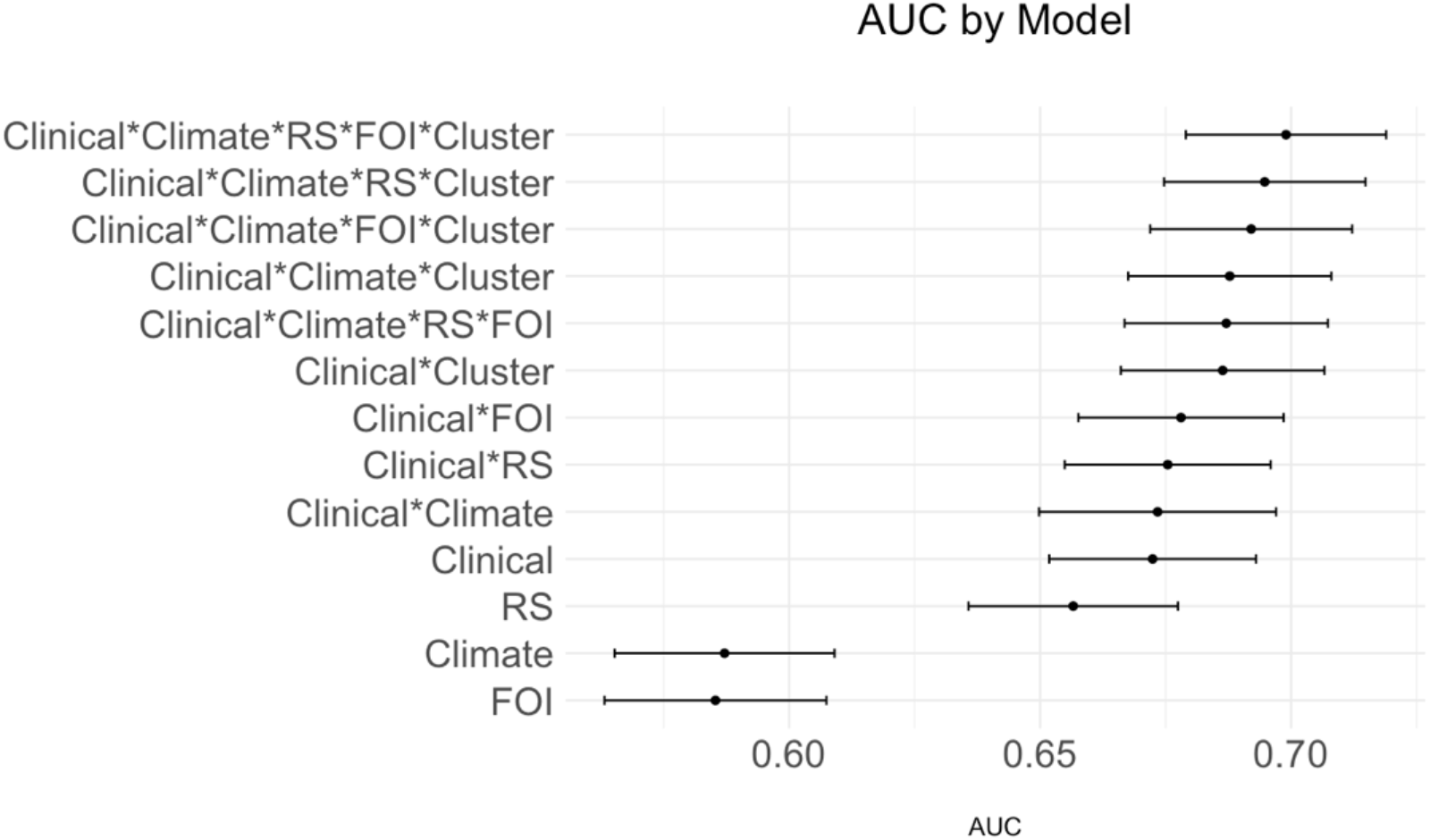
The AUCs and confidence intervals by base model, compared to base model plus inclusion of additional data sources. ‘Clinical’ indicates the inclusion of the top three clinical predictors, ‘Climate’ indicates the inclusion of climate predictors, ‘RS’ indicates the inclusion of reconstructed susceptibility estimates derived using national surveillance data, ‘FOI’ indicates the inclusion of force of infection estimates derived using cohort data, ‘Cluster’ indicates the recent case cluster metric.

### Addition of reconstructed susceptibility (RS) estimates to the clinical parameters model resulted in an improved area under the curve

Using historical hospital case data from the province, we obtained estimates of the size of the susceptible population by age for each year (across all subdistricts in the province). In our predictive model we used the prior year’s RS estimates. Using logistic regression, we found secondary RS estimates performed better than primary RS estimates [60.7% (95%CI: 58.6-62.9) vs 52.3% (95%CI:50.1-54.6)]. When added to a random forest classifier with climate and/or clinical predictors, the inclusion of RS estimates consistently resulted in higher AUCs (Table 2). When added to the top 3 clinical parameters alone, RS estimates non-significantly improved model performance from an AUC of 67.0% (95%CI: 65.0-68.8) to an AUC of 67.5% (95%CI: 65.4-69.5), (median p=0.40, 9% p-values < .05). Finally, a model including all predictors resulted in higher AUCs than a model without RS (median p=0.09, 32% p-values < 0.05).

### Addition of subdistrict-specific Force of Infection (FoI) estimates to the clinical parameters model resulted in an improved area under the curve

We incorporated FoI estimates for each age by subdistrict using data from a local cohort study. This assumes that the underlying differences in the force of infection are constant in time. Using logistic regression, FoI estimates had an AUC of 57.0% (95%CI: 54.8-59.2). The inclusion of FoI estimates lead to increases in AUC when added to the top clinical predictors, when added to clinical predictors and climate data, and when added to clinical predictors, climate predictors, and RS estimates (Table 2). When included with all other predictors, a model with FoI estimates non-significantly improved performance compared to a model without FoI estimates (median p=0.30, 23% p-values < 0.05)

### Addition of the case clustering metric to the clinical parameters model resulted in an improved area under the curve

Finally, we fit a model that assessed for clustering of recent cases based on prior patients presenting to the KPP hospital. Using logistic regression, we found the case clustering metric (the number of positive cases in the subdistrict over last 30 days divided by the total number of cases from that subdistrict in the study period) had an AUC of 56.4% (95%CI: 54.2-58.6). We found that the use of the case clustering metric consistently improved model performance. Stratifying by the finer spatial size of subdistrict consistently outperformed models with prior patients stratified by province. When added to the top performing clinical variables, model performance significantly improved (median p= 0.02, 60% of p-values < 0.05). When compared to a model with all predictors except cluster of recent cases, the inclusion of this predictor significantly improved model performance (median p= 0.007, 79% p-values <0.05).

Finally, when comparing a model including all predictors with a model including only the top clinical predictors model performance improved from an AUC of 67.0% (95%CI: 65.0-69.1) to an AUC of 70.0% [(95%CI: 67.9-71.9) (median p=0.006, 87% p-values < 0.05)].

## Discussion

The management of AFI in LMICs often requires clinical decision making with limited availability of diagnostic testing. The differential diagnosis of AFI is broad and clinicians must decide on appropriate use of antibiotics as well as patient disposition. If diagnostics are available, clinicians must consider if the benefits of the information obtained outweighs the cost of the test. CDSSs can augment clinical decision making at minimal cost to the clinician and have proven effective at improving therapeutic management and reducing unnecessary diagnostic tests in LMIC settings (*12-14*). Historically, CDSSs use only clinical and demographic information from the presenting patient. Here, we present a predictive model for DENV infection that integrates multiple sources of information both intrinsic and extrinsic to the patient, including climate data, clinical data, seroprevalence-based susceptibility estimates, and historical information from prior patients, which results in improved predictive performance.

DENV transmission can exhibit significant temporal and geographical heterogeneity even at fine spatial scales, with variations observed even among neighboring villages (*27, 35, 36*). We thus used patient-extrinsic (location-specific) data sources in our models. Although modest, the improvement in model performance with finer spatial units suggests that population-level spatial heterogeneity exists at the district level and can be applied to individual-level clinical prediction. We expect further improvements in predictive performance if finer-scale location became routinely available for case data, such as to the community level. The improvement with the use of either the province or district level case clustering metric highlights the utility of temporal predictors in clinical prediction DENV models. We also show that reconstructed susceptibility estimates, which reflect the transmission dynamics of disease and the susceptible proportion of a population, improve individual level clinical prediction on their own. Given that reconstructed susceptibility estimates may be more difficult to obtain across different settings, we favor use of the other location-specific data sources. Moreover, reconstructed susceptibility estimates may not serve as a reliable indicator of protection against DENV, as they represent a mixed concept – immunity may reflect protection due to herd immunity or may indicate increased risk of dengue infection, as higher levels of immunity may reflect higher viral circulation of the multiple DENV serotypes with significant immunologic cross-reactivity.

Transmission of DENV occurs in a seasonal pattern, and several climate variables have been found to increase DENV transmission and/or vector populations (*17-19, 28, 29*). We found visibility and relative humidity 3 months prior to presentation to be the most important predictors of DENV infection in Kamphaeng Phet, Thailand. Our findings suggest that site-specific climate variables aid in site-specific models to predict DENV infection. Appropriate lag times would need to be tuned to different sites. For use in a clinical decision support tool, the most recent climate variables could be gathered from online weather sources, based on smartphone-based detection of GPS location. An optimal utilization of this model would be through a smartphone application, as there is a scarcity of electronic medical record availability in LMICs. This would necessitate access to a smart phone device and internet connection; however, clinicians and frontline healthcare workers increasingly have access to smartphone devices, even in remote areas of LMICs (*37*).

We found the use of clinical data alone provided moderate discrimination between DENV-positive and DENV-negative patients. There were significant differences between DENV-positive and -negative patients in 16 of the 22 clinical symptoms collected on presentation, consistent with features known to distinguish dengue from other illnesses (*38, 39*). To minimize clinician input requirements (*40*), we used random forest regression to identify the optimal variables to derive a parsimonious model. We were able to achieve near-optimal performance with only three clinical variables – age, nausea, and cough. It should be noted that the input of as little as one clinical variable – age – along with other predictors can provide useful clinical information (AUC 67.9%, 95%CI: 65.6-70.0), especially in cases where other symptoms cannot be easily obtained, such as in infants, and nonverbal or comatose patients.

Our study has several limitations. First, our model was constructed using data from a single center and testing was limited to patients suspected of having dengue infection, potentially hindering the model’s generalizability to a broader population. Similarly, as there was inherent heuristic bias in the patients selected for testing, the clinical components of the model reflect this specific population, meaning other important predictors of dengue infection, such as fever, were already included in the clinician’s decision making. Our results were limited to internal cross-validation; further studies for external validation are necessary. Finally, our assessment of the use of spatial dynamics in DENV transmission was limited as cases were only matched to each district rather than sub-district or village. In the future, models that integrate cases based on a finer spatial scale may better assess the role of a patient’s residing location in prediction. Despite these limitations, we demonstrate that predictive models that include patient-extrinsic location-specific elements can improve prediction and allow for parsimonious models that minimize clinician input and should be considered in future work on clinical prediction and decision support tools.

## Data Availability

All data produced in the present study are available upon reasonable request to the authors. All data will be made available online at time of publication.

## Acknowledgments

Research reported in this publication was supported by the United States National Institutes of Health under award number R01AI135114 (to DTL), K24AI166087 (to DTL), and P01AI034533 (to ALR and KBA), the Military Infectious Disease Research Program (MIDRP), and the European Research Council (No. 804744, to HS). RJW is funded by the National Institute of Health, through Utah Stimulating Access to Research in Residency (StARR) under award R38HL143605.

Material has been reviewed by the Walter Reed Army Institute of Research. There is no objection to its presentation and/or publication. The opinions or assertions contained herein are the private views of the author, and are not to be construed as official, or as reflecting true views of the Department of the Army or the Department of Defense. The investigators have adhered to the policies for protection of human subjects as prescribed in AR 70-25.

## Conflicts of Interest

The authors have declared no conflicts of interest.

## Data Availability

De-identified data and statistical code will be available at time of publication.

---

**Supplementary Table S1.** TRIPOD checklist.

| Section/Topic | Item | Checklist Item | Page |
| --- | --- | --- | --- |
| **Title and abstract** |  |  |  |
| Title | 1 | Identify the study as developing and/or validating a multivariable prediction model, the target population, and the outcome to be predicted. | 1 |
| Abstract | 2 | Provide a summary of objectives, study design, setting, participants, sample size, predictors, outcome, statistical analysis, results, and conclusions. | 2 |
| **Introduction** |  |  |  |
| Background and objectives | 3a | Explain the medical context (including whether diagnostic or prognostic) and rationale for developing or validating the multivariable prediction model, including references to existing models. | 3 |
|  | 3b | Specify the objectives, including whether the study describes the development or validation of the model or both. | 3-4 |
| **Methods** |  |  |  |
| Source of data | 4a | Describe the study design or source of data (e.g., randomized trial, cohort, or registry data), separately for the development and validation data sets, if applicable. | 4-5 |
|  | 4b | Specify the key study dates, including start of accrual; end of accrual; and, if applicable, end of follow-up. | 4-5 |
| Participants | 5a | Specify key elements of the study setting (e.g., primary care, secondary care, general population) including number and location of centres. | 4 |
|  | 5b | Describe eligibility criteria for participants. | 4 |
|  | 5c | Give details of treatments received, if relevant. | N/A |
| Outcome | 6a | Clearly define the outcome that is predicted by the prediction model, including how and when assessed. | 4 |
|  | 6b | Report any actions to blind assessment of the outcome to be predicted. | N/A |
| Predictors | 7a | Clearly define all predictors used in developing or validating the multivariable prediction model, including how and when they were measured. | 4-6 |
|  | 7b | Report any actions to blind assessment of predictors for the outcome and other predictors. | N/A |
| Sample size | 8 | Explain how the study size was arrived at. | 4 |
| Missing data | 9 | Describe how missing data were handled (e.g., complete-case analysis, single imputation, multiple imputation) with details of any imputation method. | 6 |
| Statistical analysis methods | 10a | Describe how predictors were handled in the analyses. | 6 |
|  | 10b | Specify type of model, all model-building procedures (including any predictor selection), and method for internal validation. | 6 |
|  | 10d | Specify all measures used to assess model performance and, if relevant, to compare multiple models. | 6 |
| Risk groups | 11 | Provide details on how risk groups were created, if done. | N/A |
| **Results** |  |  |  |
| Participants | 13a | Describe the flow of participants through the study, including the number of participants with and without the outcome and, if applicable, a summary of the follow-up time. A diagram may be helpful. | 7 |
|  | 13b | Describe the characteristics of the participants (basic demographics, clinical features, available predictors), including the number of participants with missing data for predictors and outcome. | 7 |
| Model development | 14a | Specify the number of participants and outcome events in each analysis. | 7 |
|  | 14b | If done, report the unadjusted association between each candidate predictor and outcome. | N/A |
| Model specification | 15a | Present the full prediction model to allow predictions for individuals (i.e., all regression coefficients, and model intercept or baseline survival at a given time point). | 9-13 |
|  | 15b | Explain how to use the prediction model. | 9-13 |
| Model performance | 16 | Report performance measures (with CIs) for the prediction model. | 9-13 |
| **Discussion** |  |  |  |
| Limitations | 18 | Discuss any limitations of the study (such as nonrepresentative sample, few events per predictor, missing data). | 15 |
| Interpretation | 19b | Give an overall interpretation of the results, considering objectives, limitations, and results from similar studies, and other relevant evidence. | 14-15 |
| Implications | 20 | Discuss the potential clinical use of the model and implications for future research. | 14-15 |

**Supplementary Table S1. TRIPOD checklist.**
| Other information |  |  |  |
| --- | --- | --- | --- |
| Supplementary information | 21 | Provide information about the availability of supplementary resources, such as study protocol, Web calculator, and data sets. | 19-22 |
| Funding | 22 | Give the source of funding and the role of the funders for the present study. | 15 |

**Supplementary Table S2.** The relative frequencies, odds ratios, and confidence intervals for each clinical variable by DENV positivity.

| Clinical Predictors | DENV Negative | DENV Positive | OR | 95% CI |
| --- | --- | --- | --- | --- |
| Fever |  |  | 2.15 | 1.87-2.47 |
| No | 971 (14%) | 391 (6.8%) |  |  |
| Yes | 6,127 (86%) | 5,337 (93%) |  |  |
| Nausea |  |  | 1.66 | 1.53-1.79 |
| No | 4,044 (57%) | 2,543 (44%) |  |  |
| Yes | 3,054 (43%) | 3,185 (56%) |  |  |
| Headache |  |  | 1.53 | 1.4-1.67 |
| No | 2,304 (32%) | 1,371 (24%) |  |  |
| Yes | 4,794 (68%) | 4,357 (76%) |  |  |
| Emesis |  |  | 1.50 | 1.39-1.62 |
| No | 4,126 (58%) | 2,754 (48%) |  |  |
| Yes | 2,972 (42%) | 2,974 (52%) |  |  |
| Malaise |  |  | 1.33 | 1.23-1.44 |
| No | 3,687 (52%) | 2,569 (45%) |  |  |
| Yes | 3,411 (48%) | 3,159 (55%) |  |  |
| Anorexia |  |  | 1.33 | 1.22-1.44 |
| No | 4,611 (65%) | 3,340 (58%) |  |  |
| Yes | 2,487 (35%) | 2,388 (42%) |  |  |
| Abdominal Pain |  |  | 1.27 | 1.17-1.38 |
| No | 4,815 (68%) | 3,570 (62%) |  |  |
| Yes | 2,283 (32%) | 2,158 (38%) |  |  |
| Myalgias |  |  | 1.26 | 1.16-1.36 |
| No | 3,638 (51%) | 2,607 (46%) |  |  |
| Yes | 3,460 (49%) | 3,121 (54%) |  |  |
| Chills |  |  | 1.20 | 1.11-1.3 |
| No | 4,139 (58%) | 3,078 (54%) |  |  |
| Yes | 2,959 (42%) | 2,650 (46%) |  |  |
| Retro-orbital Pain |  |  | 1.16 | 1.06-1.28 |
| No | 5,506 (78%) | 4,284 (75%) |  |  |
| Yes | 1,592 (22%) | 1,444 (25%) |  |  |
| Hemorrhage |  |  | 1.16 | 1.05-1.29 |
| No | 5,951 (84%) | 4,677 (82%) |  |  |
| Yes | 1,147 (16%) | 1,051 (18%) |  |  |
| Diarrhea |  |  | 1.06 | 0.97-1.16 |
| No | 5,401 (76%) | 4,297 (75%) |  |  |
| Yes | 1,697 (24%) | 1,431 (25%) |  |  |
| Arthralgias |  |  | 1.05 | 0.96-1.15 |
| No | 5,198 (73%) | 4,134 (72%) |  |  |
| Yes | 1,900 (27%) | 1,594 (28%) |  |  |
| Rash |  |  | 1.02 | 0.93-1.12 |
| No | 5,507 (78%) | 4,426 (77%) |  |  |
| Yes | 1,591 (22%) | 1,302 (23%) |  |  |
| Age |  |  | 0.98 | 0.98-0.98 |
|  | 23 (18) | 18 (11) |  |  |
| Dark Urine |  |  | 0.86 | 0.75-1 |
| No | 6,502 (92%) | 5,306 (93%) |  |  |
| Yes | 596 (8.4%) | 422 (7.4%) |  |  |
| Seizure |  |  | 0.83 | 0.69-0.99 |
| No | 6,704 (94%) | 5,461 (95%) |  |  |
| Yes | 394 (5.6%) | 267 (4.7%) |  |  |
| Abnormal Movement |  |  | 0.79 | 0.67-0.93 |
| No | 6,622 (93%) | 5,423 (95%) |  |  |
| Yes | 476 (6.7%) | 305 (5.3%) |  |  |
| Nuchal Rigidity |  |  | 0.77 | 0.63-0.94 |
| No | 6,764 (95%) | 5,516 (96%) |  |  |
| Yes | 334 (4.7%) | 212 (3.7%) |  |  |
| Pharyngitis |  |  | 0.76 | 0.7-0.83 |
| No | 4,978 (70%) | 4,322 (75%) |  |  |
| Yes | 2,120 (30%) | 1,406 (25%) |  |  |
| Jaundice |  |  | 0.63 | 0.51-0.78 |
| No | 6,761 (95%) | 5,552 (97%) |  |  |
| Yes | 337 (4.7%) | 176 (3.1%) |  |  |
| Cough |  |  | 0.55 | 0.51-0.6 |
| No | 4,039 (57%) | 4,044 (71%) |  |  |
| Yes | 3,059 (43%) | 1,684 (29%) |  |  |
| Rhinitis |  |  | 0.55 | 0.49-0.61 |
| No | 5,641 (79%) | 5,017 (88%) |  |  |
| Yes | 1,457 (21%) | 711 (12%) |  |  |

**Supplementary Figure S1.**
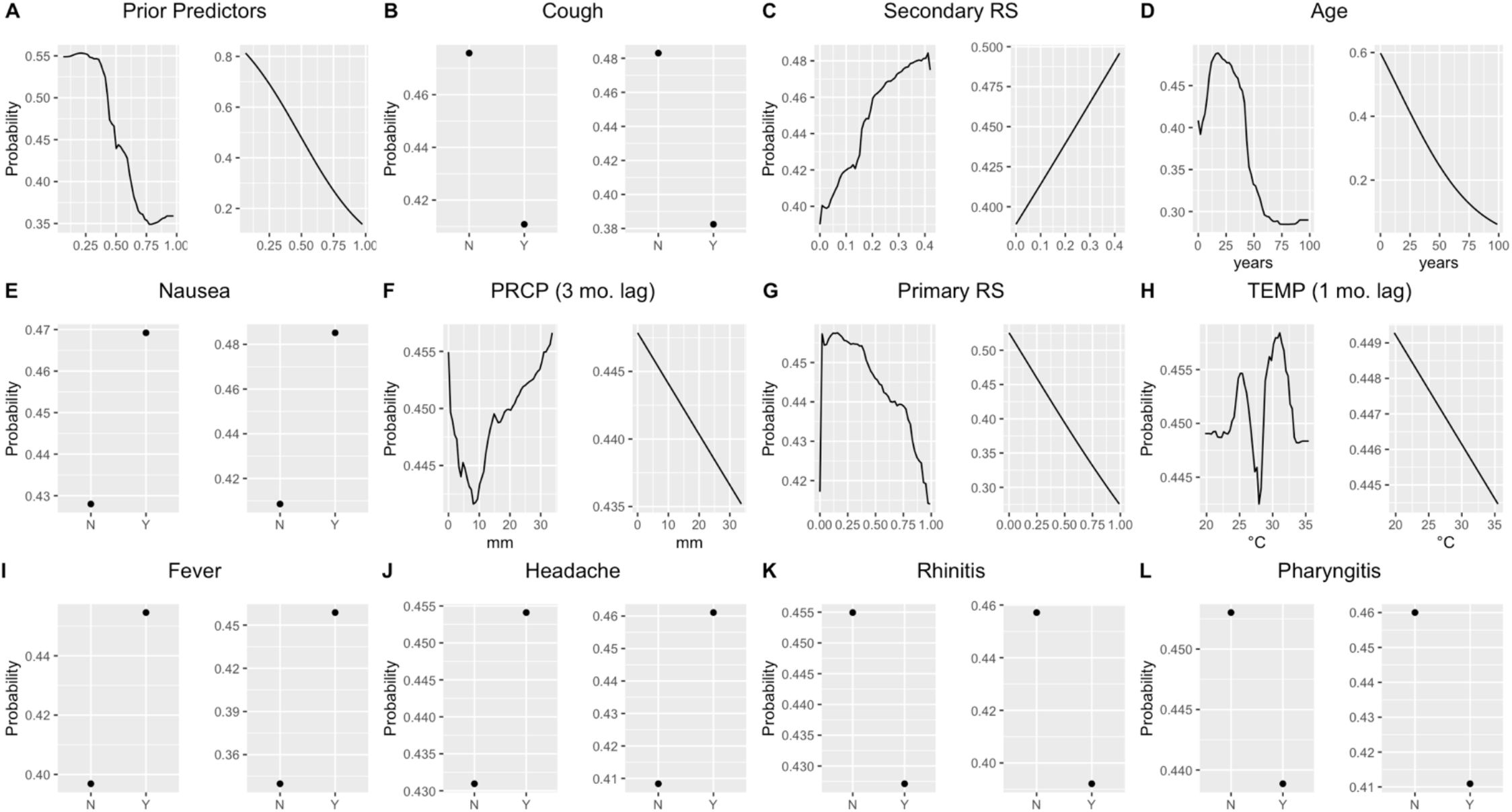
Partial Dependency Plots for the top performing variables for predicting DENV infection by AUC. For each predictor, the graph on the left shows the partial dependency for a random forest model and the partial dependency for a logistic regression model is shown on the right. ‘Y’ indicates presence of the symptom and ‘N’ indicates absence of a symptom. ‘PRCP’ refers to precipitation, ‘TEMP’ refers to the environmental temperature, ‘RS’ refers to reconstructed susceptibility estimates.

**Supplementary Table S3.** The relative frequency of the top performing clinical variables stratified by age group. ‘Y’ indicates presence of the symptom and ‘N’ indicates absence of a symptom.

|  | Overall, N<br>= 12,826<sup>1</sup> | 0-4 years,<br>N = 954<sup>1</sup> | 5-9 years,<br>N = 2,033<sup>1</sup> | 10-14<br>years, N<br>= 2,971<sup>1</sup> | 15-19<br>years,<br>N = 2,271<sup>1</sup> | 20-24<br>years,<br>N = 1,174<sup>1</sup> | 25-29<br>years,<br>N = 875<sup>1</sup> | 30-34<br>years,<br>N = 624<sup>1</sup> | 35-39<br>years,<br>N = 448<sup>1</sup> | 40+ years,<br>N = 1,476<sup>1</sup> | p-value<sup>2</sup> |
| --- | --- | --- | --- | --- | --- | --- | --- | --- | --- | --- | --- |
| **Nausea** |  |  |  |  |  |  |  |  |  |  | <0.001 |
| Y | 6,239 (49) | 341 (36) | 952 (47) | 1,515<br>(51) | 1,239 (55) | 646 (55) | 447 (51) | 320 (51) | 208 (46) | 571 (39) |  |
| **Cough** |  |  |  |  |  |  |  |  |  |  | <0.001 |
| Y | 4,743 (37) | 514 (54) | 840 (41) | 1,034<br>(35) | 790 (35) | 413 (35) | 282 (32) | 212 (34) | 146 (33) | 512 (35) |  |

<sup>1</sup>n (%)
<sup>2</sup>Pearson's Chi-squared test

**Supplementary Table S4.** The mean, standard deviation, odds ratio, and 95% CI intervals for each climate predictor.

| Climate Predictors<br>(months lagged) | DENV Negative<br>Mean (sd) | DENV Positive<br>Mean (sd) | OR | 95% CI |
| --- | --- | --- | --- | --- |
| DEWPT | 23.3°C (2.2) | 23.7°C (1.8) | 1.10 | 1.08-1.12 |
| TEMP (1) | 28.4°C (1.7) | 28.6°C (1.5) | 1.08 | 1.05-1.11 |
| DEWPT (1) | 23.3°C (2.3) | 23.6°C (1.9) | 1.08 | 1.06-1.1 |
| VISIB | 9.3 km (2.4) | 9.5 km (2.1) | 1.05 | 1.03-1.06 |
| TEMP | 28.3°C (1.6) | 28.4°C (1.4) | 1.04 | 1.01-1.06 |
| PRCP | 4.9 mm (4.5) | 5.4 mm (4.6) | 1.02 | 1.02-1.03 |
| RH | 75.3 (9.0) | 76.6 (8.1) | 1.02 | 1.01-1.02 |
| RH (3) | 70.7 (9.3) | 69.8 (8.8) | 0.99 | 0.98-0.99 |
| PRCP (3) | 3.7 mm (4.1) | 3.2 mm (3.9) | 0.97 | 0.96-0.98 |
| SLP (1) | 1008.1 mbar (2.9) | 1007.8 mbar<br>(2.6) | 0.96 | 0.94-0.97 |
| SLP | 1008.1 mbar (2.9) | 1007.7 mbar<br>(2.7) | 0.94 | 0.93-0.96 |
| VISIB (3) | 8.3 km (2.8) | 7.7 km (2.9) | 0.93 | 0.92-0.94 |
| WDSP | 0.6 m/s (0.3) | 0.6 m/s (0.3) | 0.74 | 0.65-0.85 |
| WDSP (3) | 0.7 m/s (0.3) | 0.7 m/s (0.3) | 0.73 | 0.64-0.83 |

## Notes

### Competing Interest Statement

The authors have declared no competing interest.

